# Incidence and risk factors of pulmonary hypertension after venous thromboembolism: An analysis of a large healthcare database

**DOI:** 10.1101/2020.10.25.20219303

**Authors:** Pamela L. Lutsey, Kurt W. Prins, Line H. Evensen, Rob F. Walker, Joel F. Farley, Richard F. MacLehose, Alvaro Alonso, Neil A. Zakai, Thenappan Thenappan

## Abstract

**Background:** Pulmonary hypertension (PH) is a devastating potential complication of pulmonary embolism (PE), a manifestation of venous thromboembolism (VTE). The incidence of and risk factors for PH in those with prior VTE is poorly characterized.

**Methods:** ICD codes from inpatient and outpatient medical claims from MarketScan administrative databases for years 2011-2018 were used were used to identify cases of VTE, comorbidities prior to the VTE event, and PH occurring subsequent to the VTE event. Cumulative incidence and hazard ratios (HR), and their 95% confidence intervals (CI), were calculated.

**Results:** The 170,021 VTE cases included in the analysis were on average (± SD) 57.5 ± 15.8 years old and 50.5% were female. A total of 5,946 PH cases accrued over an average follow-up of 1.94 years. Two years after incident VTE the cumulative incidence (95% CI) of PH was 3.5% (3.4%, 3.7%) overall. It was higher among women [3.9% (3.8%-4.1%)] than men [3.2% (3.0%-3.3%)], and among patients presenting with PE [6.2% (6.0%-6.5%)] than those presenting with deep vein thrombosis-only [1.1% (1.0%-1.2%)]. Adjusting for age and sex, risk of PH was higher among VTE patients with underlying comorbidities. The strongest associations were observed with concomitant heart failure [HR: 2.17 (1.04-2.31)], chronic pulmonary disease [2.01 (1.90-2.14)], and myocardial infarction [1.53 (1.40-1.67)].

**Conclusions:** In this large real-world population of insured people with VTE, 3.5% developed PH in the 2 years following their initial VTE event. Risk was higher among women, with increasing age, and in those with additional comorbidities at the time of the VTE event. These data provide insights into the burden of PH and risk factors for PH among VTE patients.

## Introduction

Pulmonary hypertension (PH) is the final physiologic process of a group of disparate diseases affecting the pulmonary vasculature, and has devastating consequences for both quality and quantity of life.^1-3^ PH is defined by an elevation in pulmonary artery pressures (PAP)^4^ which leads to a progressive increase in right ventricular afterload, thus putting increased demands on the right ventricle and eventually compromising cardiac output. Survivors of venous thromboembolism (VTE) – which consists of both pulmonary embolism (PE) and deep vein thrombosis (DVT) and afflicts approximately 1.1 million Americans annually^5^ – are at elevated risk of PH. VTE, more specifically PE, is associated with PH when pulmonary emboli/thrombi do not resolve, but instead obstruct major pulmonary arteries leading to increased PAP, right heart remodeling and PH. PH subsequent to VTE is classified by the World Health Organization^6^ and other medical entities^3,7^ as “group 4 PH”. It is also commonly referred to as chronic thromboembolic pulmonary hypertension (CTEPH).

The true incidence of PH following VTE is unknown.^3,8^ Prior literature has reported a range of 0.1% to 9.1% for the cumulative incidence of PH in VTE patients.^8-11^ To date, the largest such study reported an incidence of 1.3% 2 years after the PE event, and 3.3% 10 years after the PE.^8^ It included 23,329 VTE patients and 283 PH cases, and utilized data from a subset of the UK Clinical Practice Research Datalink (CPRD) which had linkage to data on hospitalizations and mortality.

Much also remains to be learned about factors which predict development of PH beyond experiencing a VTE event. Much of what we know about PH comes from specialized disease registries or from patients enrolled into RCTs. Patients who take part in registries or RCTs are different than the general PH patient profile due to selection bias and the often rigid criteria for enrollment into RCTs. In real-world populations, multiple pathologies may play a role in PH development after a VTE event; this population may have pre-existing autoimmune diseases (Type I), left heart disease (Type II), or chronic lung disease (Type III) in addition to VTE. As such, there is a clear need for real-world data to elucidate the burden of PH among VTE survivors, regardless of the exact etiology, and how this burden varies by demographic factors, VTE presentation and comorbid disease burden.

Using administrative data from the MarketScan databases we evaluated risk of PH subsequent to VTE. Specifically, we report incidence of PH after VTE, and risk factors for PH among VTE survivors.

## Methods

### Data source

This retrospective cohort analysis utilized IBM MarketScan® Commercial Claims and Encounter and Medicare Supplemental and Coordination of Benefits databases^12^ for calendar years 2011 through 2018. These data contain private-sector health data from some 350 different payers across the U.S. These plans contribute data to a central repository which is then standardized into a common data format for research and blinded so that specific plans cannot be identified. The large number of contributing payers and broad geographic coverage of plans increases the generalizability of our study results to commercially insured populations.^12^

These administrative databases contain individual-level, de-identified, HIPAA-compliant, healthcare claims information from U.S. employers, health plans, hospitals, and Medicare programs.^12^ Individual-level identifiers are used to link data across enrollment records and inpatient, outpatient, ancillary, and drug claims. The University of Minnesota Institutional Review Board deemed this research exempt from review.

### VTE cohort

We included in the present analysis individuals aged 18 to 99 with incident VTE, at least one prescription for an OAC within the 31 days before or after their first VTE claim, and ≥3 months of continuous enrolment prior to their first OAC prescription.^13^ We defined incident VTE as having at least one inpatient claim for VTE or two outpatient claims for VTE, which were 7 to 185 days apart, in any position, based on International Classification of Diseases (ICD) codes, with no evidence of prior VTE ICD code. We also required that beneficiaries were anticoagulant-naïve patients at the time of the VTE event. Among the VTE patients we then identified OAC prescriptions, using outpatient pharmaceutical claims data, by National Drug Codes indicating fills for apixaban, rivaroxaban or warfarin. Validation studies for apixaban and rivaroxaban claims have not yet been conducted. However, the validity of warfarin claims in administrative databases is excellent (sensitivity: 94%, positive predictive value (PPV): 99%).^14^ Our VTE definition is similar to that used in a recent validation study by Sanfilippo, et al., which reported a PPV of 91%.^15^ The Sanfilippo definition was also based on one inpatient or two outpatient VTE claims, and required evidence of treatment. We further classified VTE cases according to whether there were ICD codes for PE, or if the ICD codes only indicated DVT.

### PH identification

To define PH, we required at least 1 inpatient or 2 outpatient claims for PH (ICD-9-CM codes 416.0, 416.2, 416.8, 416.9 or ICD-10-CM codes I27.0, I27.2x, I27.82, I27.9, in any position). This is comparable to definitions used in prior analyses of PH in administrative data.^9,16,17^ Our definition was similar to that used in a medical record validation study by Wijeratne^9^ et al. In their study, based in Ontario, Canada, the authors identified 100 individuals with PH ICD codes, then validated the cases with hospital chart abstraction. The PPV was 97%. In a United Kingdom-based validation study of group 4 PH, which also validated cases identified by ICD codes with manual medical record review, the specificity was 99.2% and the sensitivity 85.8%.^8^ This study required PH ICD codes be present at a minimum of 3 months or later after initiating OACs for VTE treatment, in order to discriminate PH from ‘subacute’ PE. Current PH diagnostic guidelines also recommend abstaining from diagnosing PH until 90 days after the initial PE event.^3^ Therefore in the present analysis we required evidence of PH at least 90 days after the initial PE event.

### Identification of potential PH risk factors

The literature was reviewed to identify potential PH risk factors for exploration in the present analysis. To define these risk factors for analysis, we used information prior to the OAC initiation date (minimum 90 days) from all data sources in MarketScan (i.e., demographic data, inpatient, outpatient, and pharmacy claims). We identified 17 pre-specified comorbidities using validated algorithms^18,19^ (wherever possible) applied to the inpatient and outpatient data.

### Statistical analysis

The initial sample included 553,387 patients with ICD codes indicating VTE aged 18 to 99 years. The analytic sample was 432,950 once restricted to individuals ever prescribed an OAC between January 1, 2011 and December 31, 2018; 273,938 after requiring the first OAC prescription be within 31 days of the VTE ICD code date; 203,289 after requiring ≥90 days of continuous enrolment before the first OAC prescription; and 194,403 after excluding those with ICD codes for PH prior to the VTE event. After additionally required ≥90 days of follow-up post-VTE the final analytic sample was 170,021.

Descriptive characteristics are presented as means ± standard deviations (SD) and percentages. The Nelson-Aalen estimator was used to estimate the cumulative incidence and 95% confidence intervals (95% CI). Cox proportional hazards regression was used to estimate the association between potential risk factors and risk of incident PH among VTE patients. Follow-up began 90 days after the first OAC prescription was filled for VTE primary treatment in order to reduce the inclusion of sub-acute PH in our endpoint. Person-time accrued until incident PH, health plan disenrollment or the end of study follow-up. Cox proportional hazards models were adjusted for age (continuous) and sex. Data analyses were conducted in SAS 9.3 and STATA/SE 15.1.

## Results

### Cumulative incidence of PH

Our population of interest included 170,021 VTE cases who were on average (± SD) 57.5 ± 15.8 years old and 50.5% were female. The initial presentation included evidence of PE (with or without DVT) in 47.9%, and DVT-only in the remainder. A total of 5,943 PH cases accrued over an average follow-up of 1.94 ± 1.79 years (maximum follow-up: 7.50 years). Cumulative incidence of PH is reported in **Table 1**, for the entire follow-up and according to time-frames of interest, overall and stratified by sex and VTE presentation. Two years after incident VTE the cumulative incidence (95% CI) of PH was 3.5% (3.4%-3.7%) overall. It was higher among women [3.9% (3.8%-4.1%)] than men [3.2% (3.0%-3.3%)], and among patients presenting with PE [6.2% (6.0%-6.5%)] than those presenting with DVT-only [1.1% (1.0%-1.2%)].

**Table 1.** Cumulative incidence (%) and 95% confidence intervals of pulmonary hypertension in the 2 years* following incident venous thromboembolism: The MarketScan Databases 2011-2018

| **Time after VTE** | **All** | **Men** | **Women** | **PE** | **DVT** |
| --- | --- | --- | --- | --- | --- |
| 3-6 months | 0.90 (0.85, 0.95) | 0.78 (0.72-0.84) | 1.02 (0.95-1.09) | 1.65 (1.56-1.75) | 0.21 (0.18-0.24) |
| 6-9 months | 1.59 (1.53, 1.66) | 1.34 (1.26-1.43) | 1.84 (1.74-1.94) | 2.93 (2.81-3.06) | 0.38 (0.33-0.42) |
| 9-12 months | 2.09 (2.01, 2.17) | 1.77 (1.67-1.87) | 2.41 (2.29-2.52) | 3.82 (3.68-3.97) | 0.52 (0.47-0.57) |
| 2 years | 3.54 (3.43, 3.65) | 3.15 (3.00-3.30) | 3.93 (3.77-4.10) | 6.24 (6.03-6.45) | 1.11 (1.03-1.20) |
| 3 years | 4.74 (4.60, 4.89) | 4.26 (4.07-4.45) | 5.23 (5.02-5.44) | 8.19 (7.92-8.46) | 1.64 (1.53-1.77) |
| 4 years | 5.95 (5.77, 6.13) | 5.34 (5.10-5.59) | 6.55 (6.29-6.83) | 10.07 (9.74-10.42) | 2.24 (2.09-2.41) |
| 5 years | 7.24 (7.01, 7.48) | 6.53 (6.22-6.85) | 7.97 (7.63-8.32) | 12.12 (11.69-12.56) | 2.89 (2.68-3.12) |
| Overall | 10.64 (9.70, 11.68) | 9.53 (8.52-10.66) | 11.71 (10.27-13.35) | 18.22 (16.07-20.66) | 4.15 (3.69-4.66) |

\*A minimum of 90 days of follow-up post venous thromboembolism were required in order to limit capture of sub-acute pulmonary hypertension.

### Risk factors for developing PH

Cumulative incidence and hazard ratios (and 95% CI) for risk of PH according to VTE patient characteristics are presented in **Table 2**, with adjustment for age and sex. Risk of incident PH was higher in women than men [HR: 1.24 (1.17-1.31)] and increased linearly with age [HR: 1.02 (1.01-1.02) per year]. Risk for of PH was 6-fold higher among VTE patients initially presenting with evidence of PE [6.27 (5.93-6.64)] as compared to those presenting with DVT-only. Risk of incident PH was also significantly higher among individuals with all of the comorbidities explored, with the exception of autoimmune disease and HIV for which the associations were not significant. The strongest risk of developing PH was observed among patients who at the time of their VTE had concomitant heart failure [HR: 2.17 (1.04-2.31)], chronic pulmonary disease [2.01 (1.90-2.14)], and myocardial infarction [1.53 (1.40-1.67)].

**Table 2.** Characteristics of venous thromboembolism patients and risk of incident pulmonary hypertension: The MarketScan Databases 2011-2018

| VTE Patient Characteristic* | Incident PH | No Incident PH | Hazard ratio**<br>(95% CI) |
| --- | --- | --- | --- |
|  | **N=5,943** | **N=164,078** |  |
| **Demographics** |  |  |  |
| Sex |  |  |  |
| Male | 44.4 | 49.7 | 1 (ref.) |
| Female | 56 | 50 | 1.24 (1.17-1.31) |
| **Age** |  |  |  |
| Age, mean years ± SD | 15.3 | 15.8 | 1.02 (1.01-1.02) |
| 18-29 | 2.1 | 4.8 | 1 (ref.) |
| 30-39 | 4.7 | 8.6 | 1.15 (0.93-1.42) |
| 40-49 | 11.4 | 16.5 | 1.38 (1.14-1.68) |
| 50-59 | 23.3 | 25.8 | 1.82 (1.51-2.19) |
| 60-69 | 23.4 | 23.1 | 2.36 (1.96-2.83) |
| 70-79 | 18.1 | 11.6 | 3.00 (2.49-3.61) |
| 80-89 | 14.4 | 8 | 3.60 (2.98-4.35) |
| 90-99 | 2.7 | 1.7 | 3.71 (2.94-4.70) |
| **Clinical presentation of VTE** |  |  |  |
| DVT | 18.2 | 53.3 | 1 (Ref) |
| PE | 81.8 | 46.7 | 5.04 (4.72-5.38) |
| **Comorbidities** |  |  |  |
| Chronic pulmonary disease | 25.7 | 14.8 | 2.01 (1.90-2.14) |
| Hematological disorders | 18.8 | 15.1 | 1.32 (1.24-1.41) |
| Heart failure | 21.6 | 11.1 | 2.17 (2.04-2.31) |
| Hypertension | 66.3 | 54.4 | 1.52 (1.43-1.61) |
| Diabetes mellitus | 28.3 | 21.1 | 1.42 (1.34-1.50) |
| Myocardial infarction | 9.2 | 5.8 | 1.53 (1.40-1.67) |
| Ischemic stroke/TIA | 14.6 | 11.6 | 1.16 (1.08-1.25) |
| Peripheral artery disease | 15.8 | 11.8 | 1.25 (1.17-1.34) |
| Kidney disease | 13.2 | 9.2 | 1.46 (1.36-1.58) |
| Liver disease | 11.1 | 9.9 | 1.33 (1.23-1.45) |
| Malignancy | 22.8 | 22 | 1.06 (0.99-1.13) |
| Metastatic cancer | 8.9 | 9.9 | 1.17 (1.06-1.30) |
| Alcohol abuse | 1 | 1 | 1.66 (1.29-2.13) |
| Autoimmune disease | 24.9 | 34.6 | 0.96 (0.90-1.02) |
| HIV/AIDS | 0.4 | 0.7 | 1.33 (0.89-1.99) |

VTE – venous thromboembolism; PH – pulmonary hypertension;
\*% unless otherwise noted
\*\*Adjusted for age and sex

## Discussion

PH is a recognized complication of VTE, however few studies have evaluated the incidence of this adverse outcome in a population-based and prospective manner. Using data from the large MarketScan administrative databases we identified 170,021 insured VTE patients, of whom 5,934 subsequently developed PH. The cumulative incidence of PH among VTE survivors was 3.5% over 2 years of follow-up in this population. We also reported numerous risk factors for PH among VTE patients. This is the largest study we are aware of that has evaluated incidence and risk factors for PH in the context of VTE. Findings provide much needed data regarding the future burden of PH among VTE patients, and may be clinically useful for identifying VTE patients at elevated risk of PH.

### Incidence of PH among VTE survivors

In the present study the cumulative incidence of PH in the 2 years following incident VTE was 3.5%. Earlier publications have reported a range of 0.1–9.1%, over variable time-frames.^8-11^ The wide range of estimates in the prior literature has caused much speculation. Possible reasons for the range include differences in inclusion criteria of the populations studied, a paucity of early symptoms and difficulty in differentiating acute PE from symptoms of pre-existing CTEPH, whether PH diagnosis was triggered by clinical symptoms or routine screening, referral bias, and the duration of observation.^3,20^ As noted earlier, the largest prior study used data from a subset of the UK Clinical Practice Research Datalink (CPRD). It reported a cumulative incidence of 1.3% 2 years after the PE event, and 3.3% 10 years after the PE.^8^ Our cumulative incidence was somewhat higher than that observed in the CPRD, however it is important to note that we were capturing all PH and not just that CETPH. As expected, in our data PH incidence was higher among women, with increasing age, and among VTE cases initially presenting as PE

### Risk factors for PH among VTE survivors

We also prospectively evaluated numerous clinical comorbidities that may elevate VTE patients risk of developing PH. Comorbidities at the time of the VTE event were typically associated with greater PH risk. The strongest associations were seen for heart failure, chronic pulmonary disease, and myocardial infarction. It is important to keep in mind that this study considered any PH, and did not distinguish between PH groups. Given the way we defined PH, it is not possible to distinguish what form of PH the patients had. However, this does reflect clinical practice where rarely one putative cause of PH is identified, and the etiology is likely multifactorial.

### Strengths and limitations

The primary strength of this analysis is the large sample of VTE patients, and subsequently PH events, with a broad spectrum of clinical characteristics (such as may be seen in routine clinical practice). Generalizability is limited to U.S. VTE patients with health insurance. Misclassification is an important potential threat to the validity of this study, as it is with virtually all administrative data analyses. To minimize misclassification we used validated algorithms whenever possible.^15,18,19^ The algorithm we used to define VTE is very good; the PPV was 91% when validated in a different study population.^15^ Likewise, the PH definition we employed was verified in 97% of cases upon chart abstraction,^9^ and in a study of group 4 PH the sensitivity of a similar definition was 99%.^8^ However, PH symptoms are nonspecific and onset is insidious, therefore some cases were almost certainly undiagnosed, which would impact sensitivity. Nevertheless, the ‘missing’ cases reflect real-world clinical practice. Despite these limitations, the fact that the incidence rates and risk factors reported herein align with expectations provide some reassurance about our approach. Uncontrolled confounding is another important limitation, since we lacked information on relevant clinical information (e.g., size and anatomic location of thrombi). However, our results provide insight into risk factors for PH in the general VTE population. Though the MarketScan data has inherent limitations, it provides a unique opportunity to explore the incidence of PH following VTE in a real-world population. Prospective epidemiologic studies of PH have historically been exceedingly challenging due to the relative rarity of this condition. The use of administrative data for clinical research has growing support,^21-23^ including in the form of a 2020 U.S. Food and Drug Administration (FDA) statement on the value of real-world evidence in healthcare decisions.^23^

## Conclusions

In sum, we report the cumulative incidence of PH following VTE to be 3.5% at 2 years, using data from 170,021 insured VTE survivors who experienced 5,943 PH events. Furthermore, we provide prospective evidence suggesting that numerous comorbidities are associated with greater risk of PH, most notably chronic pulmonary disease, heart failure and myocardial infarction. These data enhance understanding of the burden of PH in this patient population, and may provide insights into the characteristics of VTE patients most likely to develop PH. Awareness of risk factors for PH in the context of VTE may increase the rate of diagnosis in primary and secondary care, and lead to earlier and better PH management.

## Data Availability

The licensing agreements through which we are accessing the data prohibit us from sharing the data more broadly. However both datasets are available for purchase. Information about how to purchase the data is provided in the following links:
- MarketScan: https://www.ibm.com/products/marketscan-research-databases/pricing
- Medicare 20% sample: https://www.resdac.org/cms-fee-information-research-identifiable-data

## References

1. Farber HW, Miller DP, McGoon MD, Frost AE, Benton WW, Benza RL. Predicting outcomes in pulmonary arterial hypertension based on the 6-minute walk distance. J Heart Lung Transplant. 2015;34(3):362–368.

2. Gall H, Felix JF, Schneck FK, et al. The Giessen Pulmonary Hypertension Registry: Survival in pulmonary hypertension subgroups. The Journal of Heart and Lung Transplantation. 2017;36(9):957–967.

3. Galie N, Humbert M, Vachiery JL, et al. 2015 ESC/ERS Guidelines for the Diagnosis and Treatment of Pulmonary Hypertension. Rev Esp Cardiol (Engl Ed). 2016;69(2):177.

4. Jameson JL. Harrison’s principles of internal medicine. In: Twentieth edition / ed. New York: McGraw-Hill Education,; 2018.

5. Virani SS, Alonso A, Benjamin EJ, et al. Heart Disease and Stroke Statistics-2020 Update. Circulation. 2020;0(0):CIR.0000000000000757.

6. Simonneau G, Gatzoulis MA, Adatia I, et al. Updated Clinical Classification of Pulmonary Hypertension. Journal of the American College of Cardiology. 2013;62(25, Supplement):D34–D41.

7. Galiè N, Channick RN, Frantz RP, et al. Risk stratification and medical therapy of pulmonary arterial hypertension. Eur Respir J. 2019;53(1):1801889.

8. Martinez C, Wallenhorst C, Teal S, Cohen AT, Peacock AJ. Incidence and risk factors of chronic thromboembolic pulmonary hypertension following venous thromboembolism, a population-based cohort study in England. Pulmonary Circulation. 2018;8(3).

9. Wijeratne DT, Lajkosz K, Brogly SB, et al. Increasing Incidence and Prevalence of World Health Organization Groups 1 to 4 Pulmonary Hypertension. Circulation: Cardiovascular Quality and Outcomes. 2018;11(2):e003973.

10. Hoeper MM, Humbert M, Souza R, et al. A global view of pulmonary hypertension. Lancet Respir Med. 2016;4(4):306–322.

11. Lang IM, Pesavento R, Bonderman D, Yuan JX. Risk factors and basic mechanisms of chronic thromboembolic pulmonary hypertension: a current understanding. Eur Respir J. 2013;41(2):462–468.

12. IBM Watson Health (TM). IBM MarketScan Research Databases for Health Services Researchers (White Paper). In: Corporation I, ed. Somers, NY 2018.

13. Lutsey PL, Zakai NA, MacLehose RF, et al. Risk of hospitalised bleeding in comparisons of oral anticoagulant options for the primary treatment of venous thromboembolism. British Journal of Haematology. 2019;185(5):903–911.

14. Garg RK, Glazer NL, Wiggins KL, et al. Ascertainment of warfarin and aspirin use by medical record review compared with automated pharmacy data. Pharmacoepidemiology and Drug Safety. 2011;20(3):313–316.

15. Sanfilippo KM, Wang T-F, Gage BF, Liu W, Carson KR. Improving Accuracy of International Classification of Diseases Codes for Venous Thromboembolism in Administrative Data. Thrombosis research. 2015;135(4):616–620.

16. George MG, Schieb LJ, Ayala C, Talwalkar A, Levant S. Pulmonary Hypertension Surveillance: United States, 2001 to 2010. CHEST. 2014;146(2):476–495.

17. Anand V, Roy SS, Archer SL, et al. Trends and Outcomes of Pulmonary Arterial Hypertension–Related Hospitalizations in the United States: Analysis of the Nationwide Inpatient Sample Database From 2001 Through 2012. JAMA Cardiology. 2016;1(9):1021–1029.

18. Cunningham A, Stein CM, Chung CP, Daugherty JR, Smalley WE, Ray WA. An automated database case definition for serious bleeding related to oral anticoagulant use. Pharmacoepidemiol Drug Saf. 2011;20(6):560–566.

19. Quan H, Sundararajan V, Halfon P, et al. Coding algorithms for defining comorbidities in ICD-9-CM and ICD-10 administrative data. Med Care. 2005;43(11):1130–1139.

20. Guérin L, Couturaud F, Parent F, et al. Prevalence of chronic thromboembolic pulmonary hypertension after acute pulmonary embolism. Thromb Haemost. 2014;112(09):598–605.

21. Parks AL, Redberg RF. Dabigatran compared with rivaroxaban vs warfarin—reply. JAMA Internal Medicine. 2017;177(5):744–744.

22. Corrigan-Curay J, Sacks L, Woodcock J. Real-world evidence and real-world data for evaluating drug safety and effectiveness. JAMA. 2018;320(9):867–868.

23. U.S. Food and Drug Administration. Real-World Evidence: Real-world data (RWD) and real-world evidence (RWE) are playing an increasing role in health care decisions. 2020; https://www.fda.gov/science-research/science-and-research-special-topics/real-world-evidence. Accessed September 1, 2020.

